# Metabolomic and lipidomic signatures in autosomal dominant and late-onset Alzheimer disease brains

**DOI:** 10.1101/2021.11.04.21265941

**Authors:** Brenna C Novotny, Maria Victoria Fernandez, Ciyang Wang, John P Budde, Kristy Bergmann, Abdallah Eteleeb, Joseph Bradley, Carol Webster, Curtis Ebl, Joanne Norton, Jen Gentsch, Umber Dube, Fengxian Wang, John C Morris, Randall J Bateman, Richard J Perrin, Eric McDade, Chengjie Xiong, Jasmeer Chhatwal, Dominantly Inherited Alzheimer Network Study Group, Alzheimer’s Disease Neuroimaging Initiative, Alzheimer’s Disease Metabolomics Consortium, Alison Goate, Martin Farlow, Peter Schofield, Helena Chui, Celeste M Karch, Bruno A Benitez, Carlos Cruchaga, Oscar Harari

## Abstract

The identification of multiple genetic risk factors for Alzheimer Disease (AD) provides evidence to support that many pathways contribute to AD onset and progression. However, the metabolomic and lipidomic profiles in carriers of distinct genetic risk factors are not fully understood. The metabolome can provide a direct image of dysregulated pathways in the brain, including information on treatment targets. In this study, we interrogate the metabolomic and lipidomic signatures in the AD brain, including carriers of pathogenic variants in *APP, PSEN1*, and *PSEN2* (autosomal dominant AD; ADAD), *APOE* ε4 and *TREM2* risk variant carriers, and non-carrier sporadic AD (sAD). We generated metabolomic and lipidomic data from parietal cortical tissue from 366 participants with AD pathology and 26 cognitively unimpaired controls using the Metabolon global metabolomics platform. We identified 133 metabolites associated with disease status (FDR *q*-value<0.05). In sAD brains these include tryptophan betaine (b=-0.57) and N-acetylputrescine (b=-0.14). Metabolites associated with sAD and ADAD include ergothioneine (b=-0.21 and -0.26 respectively) and serotonin (b=-0.34 and -0.58, respectively). *TREM2* and ADAD showed association with α-tocopherol (b=-0.12 and -0.12) and CDP-ethanolamine (b=-0.13 and -0.10). β-citrylglutamate levels are associated with sAD, ADAD, and TREM2 compared to controls (b=-0.15; -0.22; and -0.29, respectively). Additionally, we identified a signature of 16 metabolites that is significantly altered between genetic groups (sAD vs. control *p =* 1.05×10^-7^, ADAD vs. sAD *p* = 3.21×10^-5^) and is associated with Braak tau stage and disease duration. These data are available to the scientific community through a public web browser (http://ngi.pub/Metabolomics). Our findings were replicated in an independent cohort of 327 individuals.

## INTRODUCTION

Alzheimer disease (AD), the most common form of dementia, is a heterogeneous and complex disease neuropathologically characterized by the accumulation of amyloid (Aβ) plaques and neurofibrillary tangles in the brain. AD may develop as familial or sporadic. Recent advancements in AD diagnosis and treatment could benefit from a comprehensive multi-omic approach to studying diverse biological processes, including metabolism^1,2^. Pathological changes in AD begin decades before the diagnosis of AD^3^. Therefore, metabolomic changes linked to AD pathology could precede disease onset and be highly informative for predictive models and preventative medicine. Metabolic decline is one of the first physiological changes detected in patients with mild cognitive impairment (MCI) due to AD^4^. Changes in lipid and energy metabolism are proven hallmarks of AD, but there are also reports of impairments in neurotransmitter, urea cycle, purine, polyamine, and bile acid metabolisms^5^. Current symptomatic treatments (cholinesterase inhibitors and memantine) target deficits in neurotransmitters to minimize cognitive decline^6^. The dysregulation of sphingolipids and glycerophospholipids in blood samples from the Alzheimer Disease Neuroimaging Initiative (ADNI) and both blood and brain samples from the Baltimore Longitudinal Study of Aging (BLSA) cohorts have been previously reported^7–9^. These metabolites allowed discrimination between AD and controls with high accuracy, sensitivity, and specificity^10^. Blood and brain endophenotype scores were then generated that summarized the relative importance of each metabolite to the severity of AD pathology and disease progression. Furthermore, Stamate et al. (2019) used machine learning classifiers to demonstrate that a panel of plasma metabolites has the potential to match the area under the curve (AUC) of well-established cerebrospinal fluid (CSF) biomarkers when used to classify AD vs. healthy individuals^11^. Pathway analysis with the top 20 predictive metabolites indicated that the nitrogen pathway was overrepresented. Though much progress has been made in determining the specific metabolic changes in biospecimens from AD patients, the metabolomic landscape has yet to be fully understood.

AD is highly heritable and can be caused by autosomal dominant genetic variants in the amyloid precursor protein (*APP)*, presenilin-1 and -2 (*PSEN1* and *PSEN2*) genes, or associated with risk factors in multiple other loci including apolipoprotein E (Apo E) and triggering receptor on myeloid cells 2 (*TREM2*)^12–14^. The singularities of downstream effects of the complex AD genetic etiology are currently poorly understood. Pathogenic genetic variants in *APP*, which is cleaved into Aβ by β- and γ-secretase, cause altered production of Aβ. *PSEN1* and *PSEN2*, each crucial members of the γ-secretase complex, can carry pathogenic variants resulting in increased cleavage of APP into an Aβ isoform more prone to aggregation^15^. *TREM2* interacts with APOE, Aβ, and other lipids, mediating the recruitment of microglia to Aβ plaques^16,17^. Rare variants in the *TREM2* gene may lead to impaired microglial function, contributing to AD pathology^18^. Apo E is a critical player in lipid metabolism, transport, and homeostasis in the brain, and the *ε4* allele of the *APOE* gene is the main genetic risk factor for late-onset AD. Arnold et al. (2020) performed association analyses of 139 serum metabolites in the ADNI cohort and observed that females carrying the *APOE* ε4 allele experience more significant impairment of mitochondrial energy production than males^19^. These findings suggest that genetic risk factors contribute to AD pathology through distinct mechanisms. However, the metabolomic changes associated with AD pathology and with most genetic factors are currently unknown.

We sought to systematically investigate the metabolic signature of AD for carriers of the major AD genetic risk factors. In this study, we have interrogated the metabolomic and lipidomic signatures of carriers of pathogenic variants in *APP, PSEN1* or *PSEN2, APOE*, and *TREM2* risk variant carriers and compared their profiles to symptomatic AD (non-genetic), presymptomatic individuals with AD neuropathological change but no or minimal decline of cognition, and cognitively unimpaired controls without AD neuropathology. Our analysis uncovered common profiles altered across genetically categorized brains, and metabolites and lipids specific to the distinct genetic factors.

## MATERIALS & METHODS

### Cohorts

#### WUSM

Archived fresh-frozen post-mortem parietal cortical tissue samples were obtained from the Charles F. and Joanne Knight Alzheimer Disease Research Center Brain Bank (Knight ADRC) and the Dominantly Inherited Alzheimer Network (DIAN) at Washington University School of Medicine (WUSM). Samples were obtained with informed consent, and the study was approved by the WUSM Institutional Review Board. Data available for these samples included age at AD onset, age at death (AAD), gender, Clinical Dementia Rating® (CDR®)^20^, *APOE* and *TREM2* genotypes, ADAD variant status, and Braak stages for tau and Aβ. Samples were categorized based on neuropathological and genetic information: neuropathological diagnosis of AD and carrier of a pathogenic variant in any of the autosomal dominant genes (*APP, PSEN1, PSEN2*) (autosomal dominant AD [ADAD], n=25), carriers of *TREM2* risk-variants (TREM2, n=21), no known pathogenic variants (sAD, n=305), no clinical symptoms (Presymptomatic, n=15), and brains with no or minimal neuropathological AD lesions identified through post-mortem neurological examination (controls [CO], n=26) **(Table 1)**. These cohorts have been described previously^21–29^. One participant in the control group showed elevated tau pathology (Braak tau stage IV) but was classified as a control due to the absence of dementia (CDR 0) and lack of amyloid pathology (Braak Aβ stage A). This individual’s pathology is attributed to primary age-related tauopathy (PART)^30,31^.

**Table 1.**
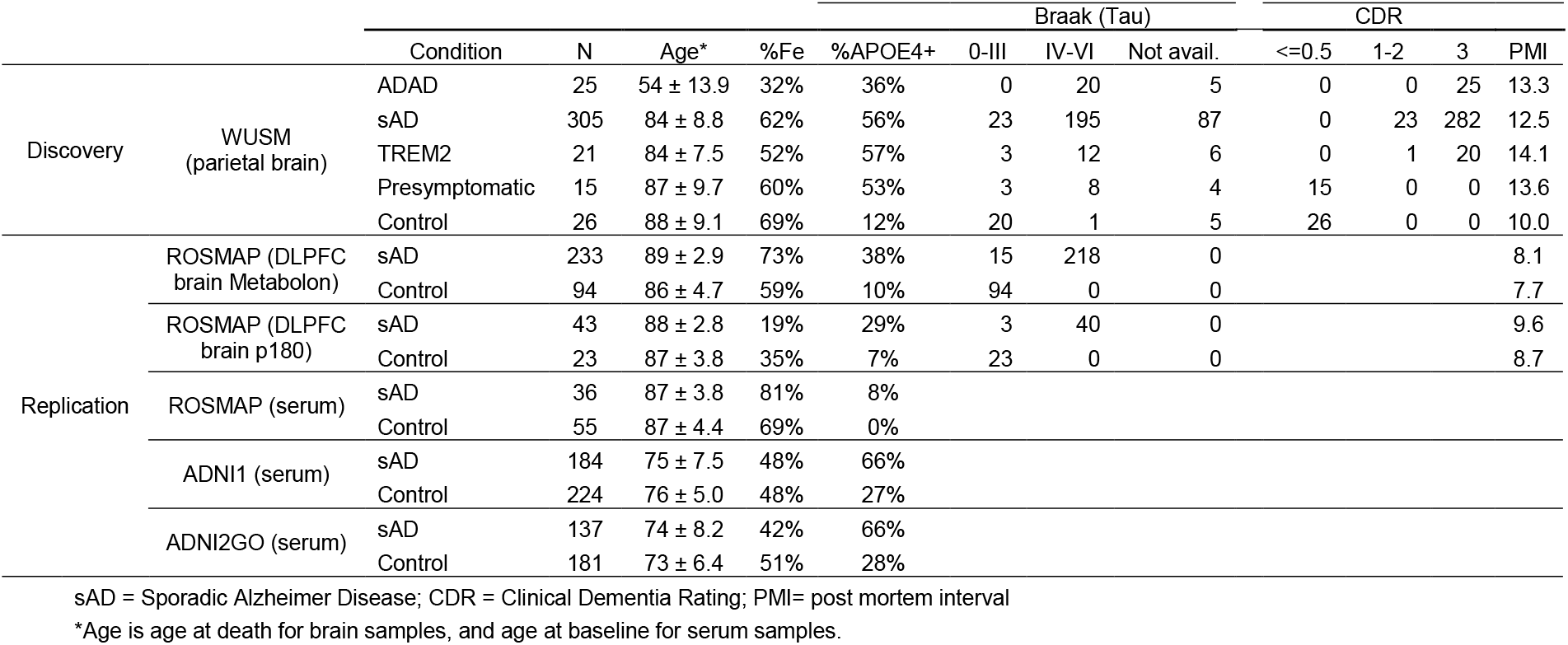
Summary statistics for the six datasets included in this study

#### ROSMAP

Data was generated by the Duke Metabolomics and Proteomics Shared Resource, a member of the ADMC, using protocols published previously for blood samples^19,32,33^; a custom protocol developed for the brain samples can be found on Synapse at syn10235609. Serum (syn10235596) and DLPFC (syn10235595) data from ROSMAP quantified on the Biocrates AbsoluteIDQ p180 platform were downloaded from Synapse in December 2020. The dorsolateral prefrontal cortex (DLPFC) metabolomic data from the ROSMAP studies quantified on the Metabolon Precision Metabolomics platform and preprocessed by the ADMC as described in^34^ were downloaded from Synapse in July 2021 (syn25878459). Details of the ROSMAP study design and methods have been described previously^35,36^. Disease status was determined based on a combination of neuropathological and cognitive metrics. Sporadic AD was defined as individuals with a CERAD assessment of “definite AD” with any Braak tau stage, or “probable AD” with Braak of at least IV.

Controls were defined by a CERAD of “possible AD” or “not AD” with Braak less than four. All AD individuals have a clinical consensus diagnosis of cognitive impairment as defined by Schneider et al. (2007), and controls have a consensus diagnosis of no cognitive impairment^37^. From the ROSMAP cohort, the following samples were analyzed: 36 sAD and 55 CO serum samples, 233 sAD and 94 CO DLPFC samples quantified with Metabolon, and 43 sAD and 23 CO DLPFC samples quantified with Biocrates p180 **(Table 1)**. For further analysis of the ROSMAP Metabolon cohort, an additional group of 223 AD and 154 CO participants was considered, based on consensus clinical diagnosis only **(Supplementary Table 1)**.

#### ADNI

Data was generated by the Duke Metabolomics and Proteomics Shared Resource, a member of the ADMC, using protocols published previously for blood samples^19,32,33^. The ADNI1 and ADNIGO/2 serum metabolomic data were obtained from the ADNI database (adni.loni.usc.edu)^33^ via the ADNIMERGE package v0.0.1 (packaged March 2018, accessed December 2020)^38^. The ADNI was launched in 2003 as a public-private partnership led by Principal Investigator Michael

W. Weiner, MD. ADNI aims to test whether neuroimaging can be combined with clinical assessment and other biological markers to measure the progression of mild cognitive impairment (MCI) and early Alzheimer disease (AD). Additional information for the ADNI studies is available at www.adni-info.org^39^. The samples analyzed from ADNI cohorts were as follows: 184 sAD and 224 CO serum samples from the ADNI1 cohort, and 137 sAD and 181 CO serum samples from the ADNIGO/2 cohort **(Table 1)**.

## Metabolite Quantification

### Metabolon Precision Metabolomics™ Platform

Data from the Knight ADRC, DIAN, and one ROSMAP cohort were generated on the Metabolon Precision Metabolomics platform. For the Knight ADRC and DIAN cohorts, 50mg frozen parietal cortical tissue samples were used for metabolite quantification. Thirteen duplicate samples served as technical replicates. The Metabolon Precision Metabolomics platform uses an ultrahigh performance liquid chromatography-tandem mass spectrometry (UPLC-MS/MS) system (Metabolon, Inc., Morrisville, USA). The platform measured 880 metabolites for the WUSM dataset and 1055 metabolites for the ROSMAP dataset. These metabolites are assigned to 111 pathways as classified by Metabolon, known as Sub Pathways. These Sub Pathways are themselves classified into nine Super Pathways: amino acids, carbohydrates, cofactors and vitamins, energy, lipids, nucleotides, peptides, xenobiotics, and partially characterized molecules (**Supplementary Table 2**).

#### Biocrates AbsoluteIDQ® p180 Platform

The remaining ADNI and ROSMAP datasets were quantified by the Biocrates AbsoluteIDQ p180 platform, which measures approximately 180 metabolites using a combination of ultra-high pressure liquid chromatography and flow-injection analysis coupled with mass spectrometry (Biocrates Life Science AG, Innsbruck, Austria). Of these 180 metabolites, 85 could be matched with those quantified by Metabolon based on Human Metabolome Database Identifier (HMDB ID) (**Supplementary Table 2**).

### Quality Control

#### Knight ADRC and DIAN Cohorts

The first step in the quality control process was to verify the quantification platform’s consistency by evaluating the technical replicates’ reproducibility. We chose the 154 metabolites with no missing values and a coefficient of variation greater than 0.3 to compare between replicate pairs via Pearson’s correlation. Each of the 13 replicate pairs showed a correlation above 0.9, demonstrating a high level of consistency. The replicate pairs were then averaged for downstream analysis. For each metabolite, if only one reading was missing from a replicate pair and the non-missing value was in the bottom 10% of the metabolite’s distribution, the non-missing value was kept. This method assumes that the missing readings in such pairs were due to the metabolite level being close to the detection limit rather than due to a technical error. Single non-missing values in the top 90% of a metabolite’s distribution were dropped.

Metabolon provided annotations for 815 of the 880 metabolites quantified analytes. The 65 remaining analytes were not assigned to known structural identities and were excluded from further analyses. We identified 198 metabolites with missing readings in at least 20% of samples. Before excluding these metabolites, Fisher’s exact tests were performed to determine if any showed differential missingness between sAD and CO, ADAD and CO, or ADAD and sAD. Those that had significantly different missingness were tested using linear regression, corrected by AAD and sex, to determine whether their non-missing readings were also significantly different in those comparisons. Those metabolites that had more missing values and lower metabolite readings in one status compared to another were rescued, assuming that their missingness was driven by a biological effect rather than a technical artifact. In all, 10 metabolites were recovered using this approach: 3-methyl-2-oxobutyrate, 4-hydroxyphenylpyruvate, acetylcholine, androsterone sulfate, cysteinylglycine disulfide, gamma-glutamyl-epsilon-lysine, gamma-glutamylphenylalanine, pregnenediol sulfate, serotonin, and tryptophan betaine. The missing readings for these 10 metabolites were imputed with each metabolite’s respective minimum reading. The 188 remaining metabolites missing at least 20% of readings were excluded from the dataset (**Supplementary Figure 1)**.

Raw readings were log10-transformed to better approximate a normal distribution. Outlier readings (outside 1.5 x interquartile range) for each metabolite were excluded, and the mean of each metabolite’s distribution was adjusted to zero.

After the metabolite QC, 95% of samples were missing less than 5% of metabolite readings; the maximum missingness for a sample was 11%. No samples were excluded due to the missingness rate, and the remaining missing values were not imputed. Principal component analysis was then performed on the scaled and imputed data provided by Metabolon to identify outlier samples using the R function PCA from the FactoMineR package^40^. Four outlier samples were excluded **(Supplementary Figure 2)**. The final dataset consisted of 627 metabolites measured in 392 samples **(Supplementary Table 3)**.

#### Replication Datasets

We employed the above-described procedure to perform the data cleaning and QC of the ADNI and ROSMAP datasets. Briefly, replicates were averaged, removing single readings above the tenth percentile of the metabolite’s distribution. Metabolites without assigned structural identities and metabolites missing at least 20% of readings were removed, readings were log10-transformed, outlier readings were removed, and the mean of each metabolite’s distribution was adjusted to zero. For each dataset, samples missing greater than 20% of readings were excluded: one from ROSMAP p180 brain, two from ROSMAP Metabolon brain, two from ADNI1, and one from ADNIGO/2. Five metabolites with missingness higher than 20% were recovered from the ROSMAP Metabolon cohort according to the procedure described above, considering associations with AD for both the neuropathological and clinical diagnoses: saccharopine, tryptophan betaine, memantine, retinol (Vitamin A) and 6-oxopiperidine-2-carboxylate. Missing values for these metabolites were imputed with the metabolites’ minimum readings. After QC, the ADNI1 dataset included 149 metabolites in 408 samples, the ADNIGO/2 dataset included 157 metabolites in 318 samples, the ROSMAP serum dataset included 162 metabolites in 91 samples, and the ROSMAP Metabolon brain dataset included 595 metabolites in 327 samples. The ROSMAP p180 brain dataset consisted of 157 metabolites in 66 samples (**Table 1**).

### Statistical analyses

Association analyses of metabolite abundance with disease status were conducted using linear regression in R software version 3.6^41^. Metabolite levels were modeled by disease status (sAD, ADAD, and TREM2) compared to CO, corrected by sex, AAD, and post-mortem interval (PMI). Associations with *APOE ε4* carrier status were also tested within the sAD status, corrected by the same variables. AAD and PMI were chosen as covariates in the model due to their correlation with the first principal component of the metabolites that passed QC (*p*<0.01). When comparing ADAD with CO, AAD was not included as it is collinear with ADAD status. Linear regression was also used to test each metabolite’s association with AAD in the sAD status group, correcting for sex and PMI. The false discovery rate (FDR) was controlled using Benjamini-Hochberg correction (R function p.adjust). The *q*-value threshold for significance was established as *q* < 0.05. To test differences in effect size between groups, we employed an analysis of covariance (ANCOVA) comparing the effects for metabolites in sAD and TREM2 relative to their effect in ADAD. We performed additional ANCOVA tests with individuals matched by CDR (CDR = 3), tau (Braak tau > 3), and Aβ (Braak Aβ = 3) to test whether the differences in effect sizes were influenced by neuropathology.

We calculated the first principal component of readings for metabolites which were differentially abundant in multiple groups, similar to the “eigengene” concept in the WGCNA package^42^. We tested the differences in the *eigenmetabolite* profile between groups. We also tested for association with CDR, Braak tau stage, and disease duration, defined as the difference of age at disease onset (AAO) and AAD. The linear regression models were corrected for sex, PMI, and AAD.

We conducted additional analyses in the ADNI and ROSMAP public metabolomics datasets to follow up on our results. For the analyses in serum, PMI was not used as a covariate as it was not applicable, and age at blood draw was used in the model rather than AAD. A meta-analysis was also performed combining the serum data from the ADNI1, ADNIGO/2, and ROSMAP datasets. The meta-analysis was carried out with the same linear model, pooling all 817 serum samples: 357 sAD and 460 CO. Meta-analysis models were additionally corrected by study.

A heatmap was constructed using Metabolon’s scaled and minimum-imputed metabolite readings with the ComplexHeatmap^43^ and circlize^44^ R packages. Individuals were separated into status groups (ADAD, sAD, TREM2, Presymptomatic, and CO), and the heatmap was additionally annotated with CDR and Braak tau scores for each individual. Hierarchical clustering of individuals was performed using the ward.D2 method in the Heatmap function from the ComplexHeatmap package.

Pathway and network analyses were performed using MetaboAnalyst^45,46^ and IMPaLA pathway over-representation analysis^47^. HMDB IDs for 105 of the 133 significant metabolites could be determined and input into MetaboAnalyst and IMPaLA. MetaboAnalyst matched 103 of those IDs to its database, while IMPaLA matched 74 **(Supplementary Table 4)**. Pathways were also explored in the Kyoto Encyclopedia of Genes and Genomes (KEGG) pathway database^48–50^. The source code of the scripts employed to QC, clean and analyze the data is available at http://github.com/HarariLab/Metabolomics.

### Pharmacological Analysis

We obtained pharmacological data for the research participants to determine any potential confounding effects of medications in our analyses. Longitudinal pharmacological data were available for 297 research participants from the Knight ADRC cohort. The number of time points (clinical assessment dates) for each participant ranged from one to 20 visits, with a mean of 3.9. The mean number of years between the most recent clinical assessment and year of death was 2.8 ± 2.5, with a range of 0-14 years. Drug group information and alternate medication names were obtained from the KEGG Drug database^48–50^. To test for confounding medication effects, we performed linear regression to test for association between medications and potentially affected metabolites. We also repeated the association analyses, including only individuals who had not been administered the medications within the five years preceding their death, to confirm that the differential abundance was not influenced by medications.

### Web Browser

Our results are available through a public browser at http://ngi.pub/Metabolomics. The browser was created using R Shiny version 1.4.0^51^ and the shinydashboard^52^, shinydashboardPlus^53^, plotly^54^, DT^55^, shinyjs^56^, htmlwidgets^57^, RColorBrewer^58^, kableExtra^59^, and dplyr^60^ R packages. Source code is publicly available in the GitHub repository http://github.com/HarariLab/Metabolomics.

## RESULTS

### Study Design

In this study, we performed a metabolomics analysis of parietal cortical tissue from participants of the DIAN and Knight ADRC cohorts (**Figure 1**). We determined the metabolomic profile of 392 participants, including three AD genetic subgroups: autosomal dominant AD (ADAD), carriers of risk variants in *TREM2* (TREM2), and sporadic AD (sAD). Detailed phenotypic information included genetic risk factors, CDR, and Braak staging for tau and Aβ. Out of the 880 metabolites quantified, 627 passed quality control (**Supplementary Table 3**). We tested differential abundance using linear models adjusted for AAD, sex, and PMI and compiled the differentially abundant metabolites into a profile to distinguish between AD genetic groups. We also conducted a pathway analysis with the significantly associated metabolites. To validate our results further, we performed association analysis on a total of 393 brain samples and 817 serum samples from both the ROSMAP and ADNI cohorts.

**Figure 1.**
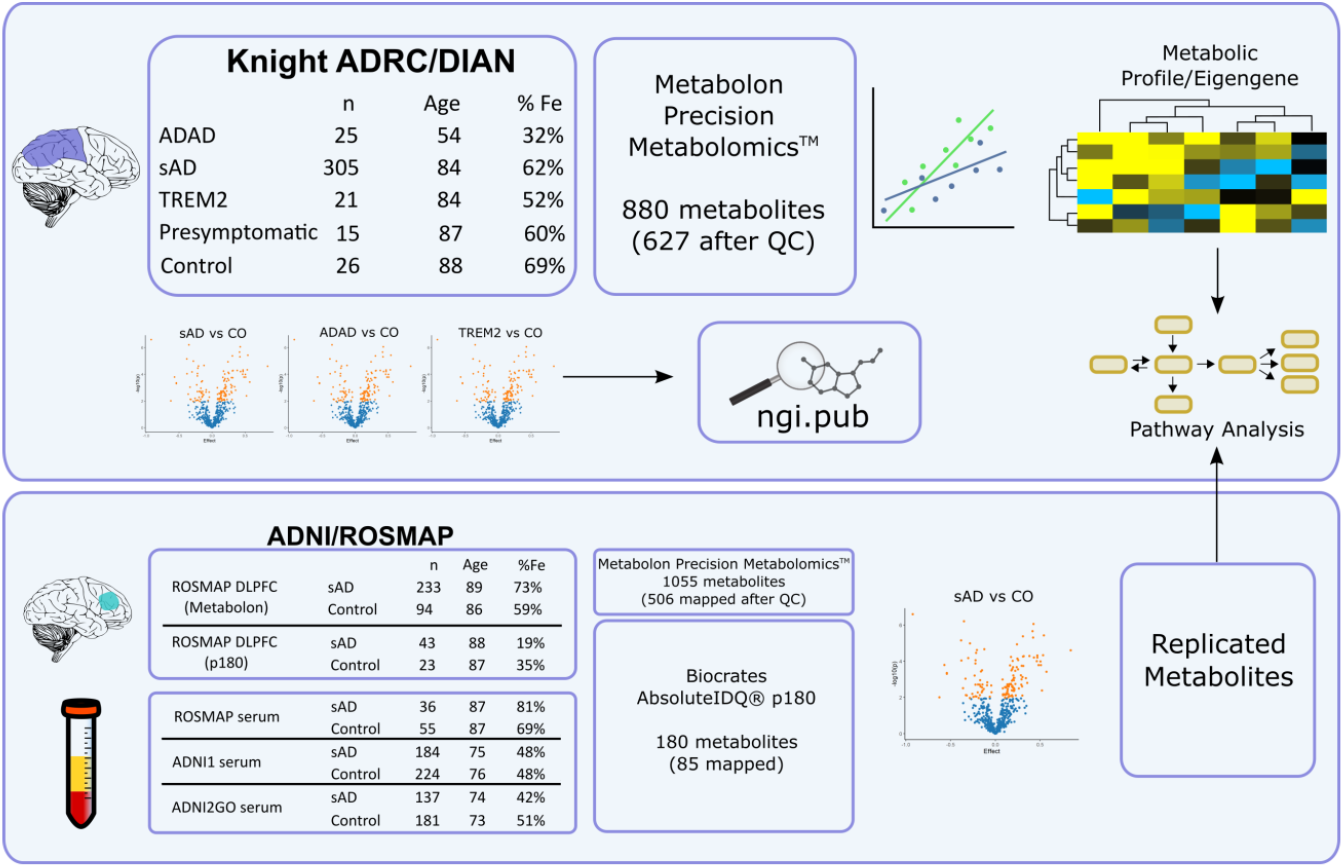
Study design. Parietal cortical tissue from donors to the Knight ADRC and DIAN were analyzed on the Metabolon Precision Metabolomics platform: autosomal dominant AD (ADAD, n=25), sporadic AD (sAD, n=305), TREM2 (n=21), Presymptomatic (n=15), and healthy control (CO, n=26). After quality control, 627 metabolites were tested for differential abundance via linear modeling. A metabolic profile was generated from 16 metabolites in common between groups. Pathway analysis was performed on the differentially abundant metabolites, and a web browser was created to share the data and results. Findings were validated in five independent datasets: dorsolateral prefrontal cortex (DLPFC) tissue from the ROSMAP cohort analyzed on the Metabolon platform (sAD n=233, CO n=94), as well as four datasets quantified using the Biocrates p180 platform: DLPFC from the ROSMAP cohort (sAD=43, CO=23), serum from the ROSMAP cohort (sAD n=36, CO n=55), serum from the ADNI1 cohort (sAD n=184, CO n=224), and serum from the ADNIGO/2 cohort (sAD n=137, CO n=181). The ROSMAP Metabolon dataset was found to have 506 metabolites in common with the Knight ADRC/DIAN cohort after quality control. The p180 platform was found to have 85 metabolites in common with the Metabolon platform.

### Metabolite association analysis identifies differential β-citrylglutamate levels in sporadic AD and ADAD

Association analysis indicated that the ADAD group had the most distinct brain metabolomics profile (with 131 significant metabolites; **Supplementary Table 5**), whereas the profiles for TREM2 and sAD cases showed a lower number of differentially abundant metabolites, with only three (α-tocopherol, β-citrylglutamate, and CDP-ethanolamine) and five (β-citrylglutamate, ergothioneine, serotonin, tryptophan betaine, and N-acetylputrescine) significant metabolites respectively **(Figure 2a-d)**. The Super Pathways represented in the ADAD-associated metabolites were Amino Acids (48 metabolites), Carbohydrates (12), Cofactors and Vitamins (9), Energy (2), Lipid (30), Nucleotide (12), Peptide (12), and Xenobiotics (6). We found that 99 of the 131 significant metabolites were independent of AAD **(Supplementary Table 6)**.

**Figure 2.**
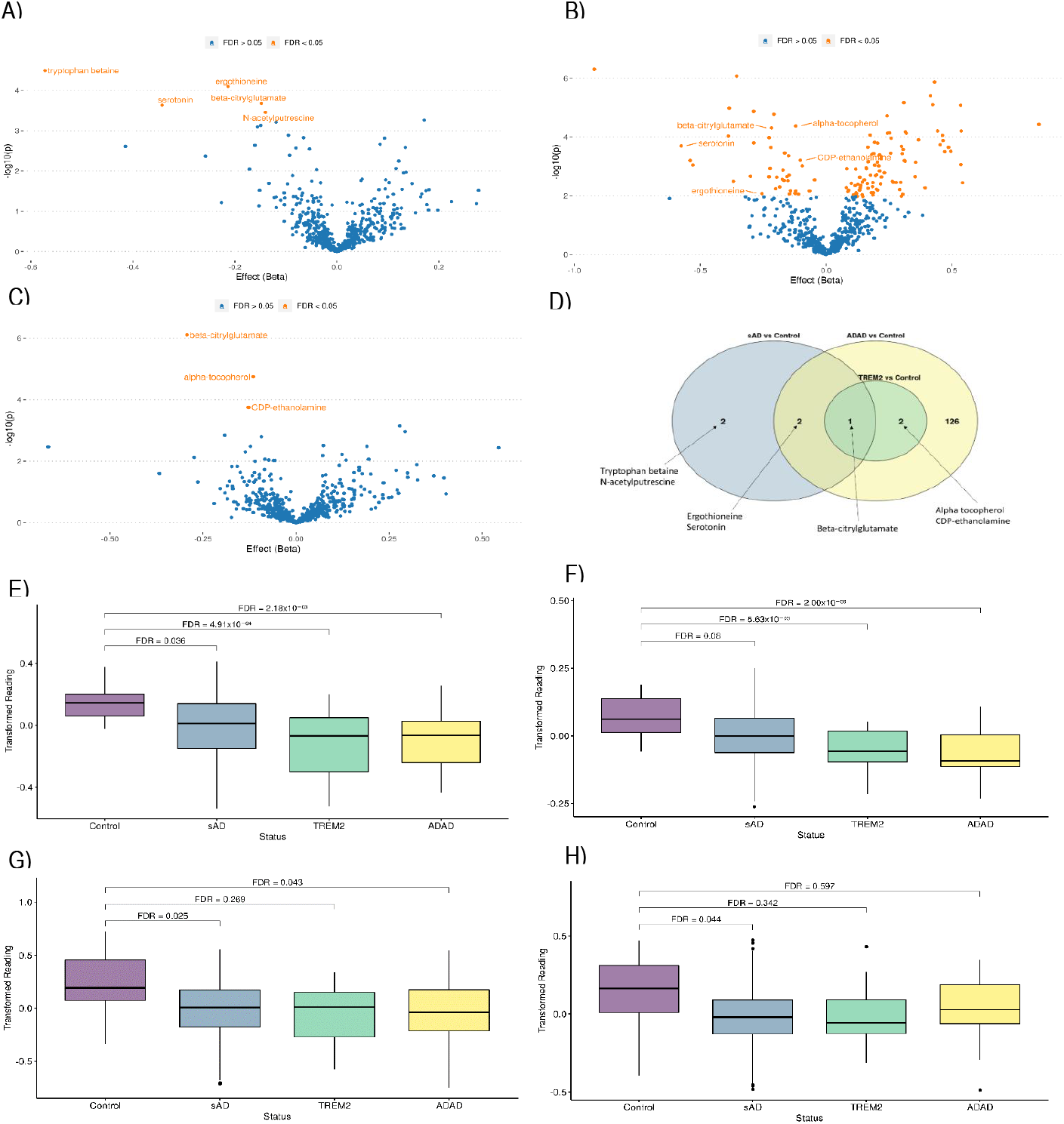
Association analysis in WUSM dataset. Volcano plots for A) sAD vs CO, B) ADAD vs CO, C) TREM2 vs CO. D) Venn diagram. Box plots for abundance of top metabolites E) β-citrylglutamate. F) α-tocopherol, G) ergothioneine, H) N-acetylputrescine.

Pathway analysis performed with these 131 metabolites indicated overrepresented pathways in the categories of amino acid metabolism, which accounted for the most pathways, as well as sphingolipid and vitamin metabolism **(Supplementary Tables 7 and 8)**. We observed perturbations in several amino acid metabolism pathways, including glutamate, glutathione, tryptophan, lysine and histidine metabolisms. Perturbations in sphingolipid metabolism have been identified previously as potential biomarkers for AD^10^. Altered amino acid metabolism has also been reported in multiple metabolomic studies of the AD brain^19,32,61^. The most notable amino acid pathways in our analysis were glutamate, glutathione, and tryptophan metabolism. Abnormal glutamate metabolism is known to cause excitotoxicity^62^, and alterations in glutathione metabolism may contribute to oxidative damage and neuronal loss^63^. Alterations in tryptophan metabolism, especially serotonin imbalances, have also been previously noted in the AD metabolome^64^. We also observed perturbations in lysine and histidine metabolism pathways and novel associations in vitamin pathways.

β-citrylglutamate (BCG) was the only metabolite significantly differentially abundant in all three genetic groups. Zhao et al. (2019) showed that serum BCG levels were significantly affected by the administration of fluoxetine, a commonly prescribed SSRI antidepressant^65^. In our study, five participants were documented as having taken fluoxetine at the time of their most recent clinical assessment. Five sAD participants and one presymptomatic participant had taken fluoxetine within the five years preceding their death **(Supplementary Table 9)**. A binomial logistic regression showed that BCG levels were not associated with fluoxetine use within the five years preceding their death in individuals with sAD (*p =* 0.98). Furthermore, we repeated the linear regression for sAD vs. CO, excluding the individuals who had taken fluoxetine in the past five years, and did not observe a change in the association of BCG levels between sAD and CO (effect = -0.15, *q* = 3.7×10^-2^). AAD was not associated with lifetime usage of fluoxetine (*p =* 0.83), nor were AAO (*p =* 0.63) or disease duration (*p =* 0.15).

### *APOE ε4* carrier status shows nominal associations with metabolites

In an association analysis of *APOE ε4* carriers vs. non-carriers in the sAD group, none of the 627 metabolites tested were significantly associated after FDR correction, though 25 metabolites were nominally significant (*p* < 0.05) (**Supplementary Table 10**).

### Follow-up in independent datasets

We tested the differential abundance of metabolites in serum and DLPFC samples from the ADNI1, ADNIGO/2, and ROSMAP cohorts to independently validate our results. For each of these cohorts, metabolites were quantified using the Biocrates AbsoluteIDQ p180 platform, which we found to have 85 metabolites in common with the Metabolon Precision Metabolomics platform. Additionally, 379 DLPFC samples were analyzed from the ROSMAP cohort, quantified on the Metabolon platform; in this dataset, 506 metabolites were in common with the Knight ADRC cohort after QC. We identified 44 metabolites that were significantly differentially abundant with consistent effect direction in our ADAD vs. CO analysis and at least one independent dataset. Among the replicated metabolites were α-tocopherol, BCG, and serotonin (**Supplementary Table 11**).

Of the seven analytes that were significant in both serum and brain cohorts, distinct direction of effect between tissue types was identified among five analytes. Specifically, 2-aminoadipate, isoleucine, valine, glutamate, and tyrosine showed a positive effect in the ADAD samples and ROSMAP sAD brains but showed a negative effect in the serum analyses. Similarly, Huo, et al. (2020) observed opposite directions of effect for glycerophospholipids between brain and serum in the ROSMAP cohort^66^. Serotonin and 1-linoleoyl-2-arachidonoyl-GPC showed concordant effects between the tissues, with lower abundance in ADAD and sAD than controls.

The replicated metabolites supported our previous pathway analysis findings. Components of nicotinamide metabolism (trigonelline), vitamin A metabolism (retinol/Vitamin A), and tocopherol metabolism (α-tocopherol/vitamin E), were replicated, supporting the role of vitamin pathways. Asparagine, methionine, threonine, and tyrosine, all part of the gamma-glutamyl cycle, were found in the replicated metabolites, along with serotonin of the tryptophan metabolism pathway. Finally, BCG, glutamate, and N-acetyl-aspartyl-glutamate were each replicated, implicating the dysregulation of glutamate metabolism.

Like the WUSM cohort, no metabolites were significant after correction when testing metabolite associations for APOE ε4 carriers vs. non-carriers. However, 23 were nominally significant (*p* < 0.05), of which none were replicated from the WUSM cohort (**Supplementary Table 10**).

### A metabolic profile associated with AD duration and Braak stage

We sought to investigate whether ADAD, TREM2, and sAD showed a similar or more distinct difference in their altered metabolomic profile. Thus, we selected the 17 metabolites that were significantly associated in the ADAD brains (*q*-value <0.05) that also were nominally associated in both the sAD and TREM2 when compared to controls (**Table 2**). Of these, ergothioneine, serotonin, BCG, CDP-ethanolamine, and α-tocopherol were statistically significant after FDR correction in sAD (ergothioneine, serotonin, and BCG) or TREM2 (BCG, CDP-ethanolamine, and α-tocopherol) (**Table 2**). These five metabolites showed lower abundance in the AD groups as compared to controls. This group of 17 metabolites was considered for the identification of a metabolic profile differentiating between status groups. Serotonin was excluded from the profile because the ADAD group was missing 17 of 25 readings for serotonin (68%). In the ROSMAP Metabolon cohort, the differential abundance of nine out of the 16 remaining metabolites was replicated after FDR correction: γ-glutamylthreonine, β-citrylglutamate, glutamate, N-acetylglutamate, 1,5-anhydroglucitol, glutarate, CDP-choline, retinol, and α-tocopherol. Additionally, aspartate, ergothioneine, 2-methylcitrate/homocitrate, and glycerophosphoinositol were nominally significant, and 3-hydroxy-2-ethylpropionate did not pass QC.

**Table 2.** Effects, p-values, and q-values for 16 metabolites which were significant after FDR correction in the ADAD vs CO comparisons and at least nominally significant in the AD vs CO and TREM2 vs CO comparisons in the WUSM cohort. Highlighted in bold are metabolites with significant Q values in the AD vs CO or TREM2 vs CO comparisons.

| Metabolite | ADAD vs CO |  |  | sAD vs CO |  |  | TREM2 vs CO |  |  | ROSMAP (DLPFC Metabolon) |  |  |
| --- | --- | --- | --- | --- | --- | --- | --- | --- | --- | --- | --- | --- |
|  | Effect | p-value | q-value | Effect | p-value | q-value | Effect | p-value | q-value | Effect | p-value | q-value |
| aspartate | 0.160 | 1.1x10<sup>-3</sup> | 1.1x10<sup>-2</sup> | 0.085 | 2.2x10<sup>-3</sup> | 9.8x10<sup>-2</sup> | 0.123 | 6.0x10<sup>-3</sup> | 0.27 | 0.042 | 1.1x10<sup>-2</sup> | 5.4x10<sup>-2</sup> |
| γ-glutamylthreonine | 0.263 | 3.2x10<sup>-3</sup> | 2.3x10<sup>-2</sup> | 0.171 | 5.4x10<sup>-4</sup> | 5.5x10<sup>-2</sup> | 0.278 | 7.1x10<sup>-4</sup> | 0.11 | 0.095 | 5.1x10<sup>-4</sup> | **5.4x10<sup>-3</sup>** |
| β-citrylglutamate | -0.217 | 4.9x10<sup>-5</sup> | 2.2x10<sup>-3</sup> | **-0.148** | **2.1x10<sup>-4</sup>** | **3.6x10<sup>-2</sup>** | **-0.293** | **7.8x10<sup>-7</sup>** | **4.9x10<sup>-4</sup>** | -0.062 | 3.7x10<sup>-4</sup> | **4.2x10<sup>-3</sup>** |
| glutamate | 0.086 | 7.1x10<sup>-3</sup> | 3.9x10<sup>-2</sup> | 0.048 | 1.7x10<sup>-2</sup> | 0.36 | 0.071 | 6.5x10<sup>-3</sup> | 0.27 | 0.060 | 1.1x10<sup>-8</sup> | **1.4x10<sup>-6</sup>** |
| N-acetylglutamate | -0.207 | 1.7x10<sup>-5</sup> | 1.0x10<sup>-3</sup> | -0.080 | 2.7x10<sup>-3</sup> | 9.8x10<sup>-2</sup> | -0.122 | 3.2x10<sup>-3</sup> | 0.19 | -0.033 | 6.8x10<sup>-4</sup> | **6.7x10<sup>-3</sup>** |
| ergothioneine | -0.255 | 8.3x10<sup>-3</sup> | 4.3x10<sup>-2</sup> | **-0.213** | **8.0x10<sup>-5</sup>** | **2.5x10<sup>-2</sup>** | -0.274 | 7.5x10<sup>-3</sup> | 0.27 | -0.092 | 2.5x10<sup>-2</sup> | 0.10 |
| 3-hydroxy-2-ethylpropionate | 0.177 | 2.0x10<sup>-3</sup> | 1.7x10<sup>-2</sup> | 0.099 | 4.4x10<sup>-2</sup> | 0.49 | 0.195 | 8.7x10<sup>-3</sup> | 0.27 |  |  |  |
| 1,5-anhydroglucitol (1,5-AG) | 0.310 | 6.6x10<sup>-6</sup> | 7.3x10<sup>-4</sup> | 0.131 | 1.1x10<sup>-2</sup> | 0.27 | 0.155 | 4.3x10<sup>-2</sup> | 0.47 | 0.088 | 4.4x10<sup>-3</sup> | **2.8x10<sup>-2</sup>** |
| 2-methylcitrate/homocitrate | -0.171 | 1.3x10<sup>-3</sup> | 1.2x10<sup>-2</sup> | -0.087 | 1.3x10<sup>-2</sup> | 0.30 | -0.118 | 2.9x10<sup>-2</sup> | 0.41 | -0.048 | 1.1x10<sup>-2</sup> | 5.4x10<sup>-2</sup> |
| glutarate (C5-DC) | 0.252 | 7.3x10<sup>-5</sup> | 2.3x10<sup>-3</sup> | 0.121 | 5.7x10<sup>-3</sup> | 0.17 | 0.170 | 1.6x10<sup>-2</sup> | 0.35 | 0.090 | 1.5x10<sup>-5</sup> | **4.0x10<sup>-4</sup>** |
| CDP-choline | -0.095 | 9.6x10<sup>-4</sup> | 1.0x10<sup>-2</sup> | -0.040 | 4.6x10<sup>-2</sup> | 0.49 | -0.093 | 1.6x10<sup>-3</sup> | 0.14 | 0.042 | 7.5x10<sup>-3</sup> | **4.1x10<sup>-2</sup>** |
| CDP-ethanolamine | -0.102 | 6.1x10<sup>-4</sup> | 7.7x10<sup>-3</sup> | -0.054 | 2.8x10<sup>-3</sup> | 9.9x10<sup>-2</sup> | **-0.128** | **1.8x10<sup>-4</sup>** | **3.7x10<sup>-2</sup>** | -0.024 | 8.2x10<sup>-2</sup> | 0.23 |
| glycerophosphoinositol | -0.164 | 3.4x10<sup>-4</sup> | 5.3x10<sup>-3</sup> | -0.095 | 1.3x10<sup>-3</sup> | 8.0x10<sup>-2</sup> | -0.098 | 2.7x10<sup>-2</sup> | 0.41 | -0.021 | 4.5x10<sup>-2</sup> | 0.15 |
| nicotinamide (vitamin B3) | -0.066 | 6.8x10<sup>-3</sup> | 3.7x10<sup>-2</sup> | -0.032 | 1.9x10<sup>-2</sup> | 0.36 | -0.056 | 9.4x10<sup>-3</sup> | 0.27 | 0.008 | 0.33 | 0.56 |
| α-tocopherol (vitamin E) | -0.120 | 4.1x10<sup>-5</sup> | 2.0x10<sup>-3</sup> | -0.065 | 1.5x10<sup>-3</sup> | 8.0x10<sup>-2</sup> | **-0.115** | **1.8x10<sup>-5</sup>** | **5.6x10<sup>-3</sup>** | -0.123 | 3.0x10<sup>-5</sup> | **5.7x10<sup>-4</sup>** |
| retinol (Vitamin A) | -0.287 | 1.3x10<sup>-5</sup> | 9.2x10<sup>-4</sup> | -0.156 | 8.0x10<sup>-4</sup> | 5.6x10<sup>-2</sup> | -0.192 | 1.4x10<sup>-3</sup> | 0.14 | -0.196 | 2.9E-04 | **3.7x10<sup>-3</sup>** |

We next compared the magnitude of the effects of the remaining 16 common metabolites across the three genetic groups. The effect in ADAD tended to be greater than that of TREM2, which was in turn greater than the effect in sAD (**Figure 3B**). An ANCOVA test showed that the relative effects of sAD and TREM2 to ADAD were significantly different (*p =* 4.37×10^-04^). This difference in effect was reproduced when individuals were matched by CDR (*p =* 2.54×10^-02^) as well as Braak stage for Tau (*p =* 1.18×10^-03^) and Aβ (*p =* 2.20×10^-03^).

**Figure 3.**
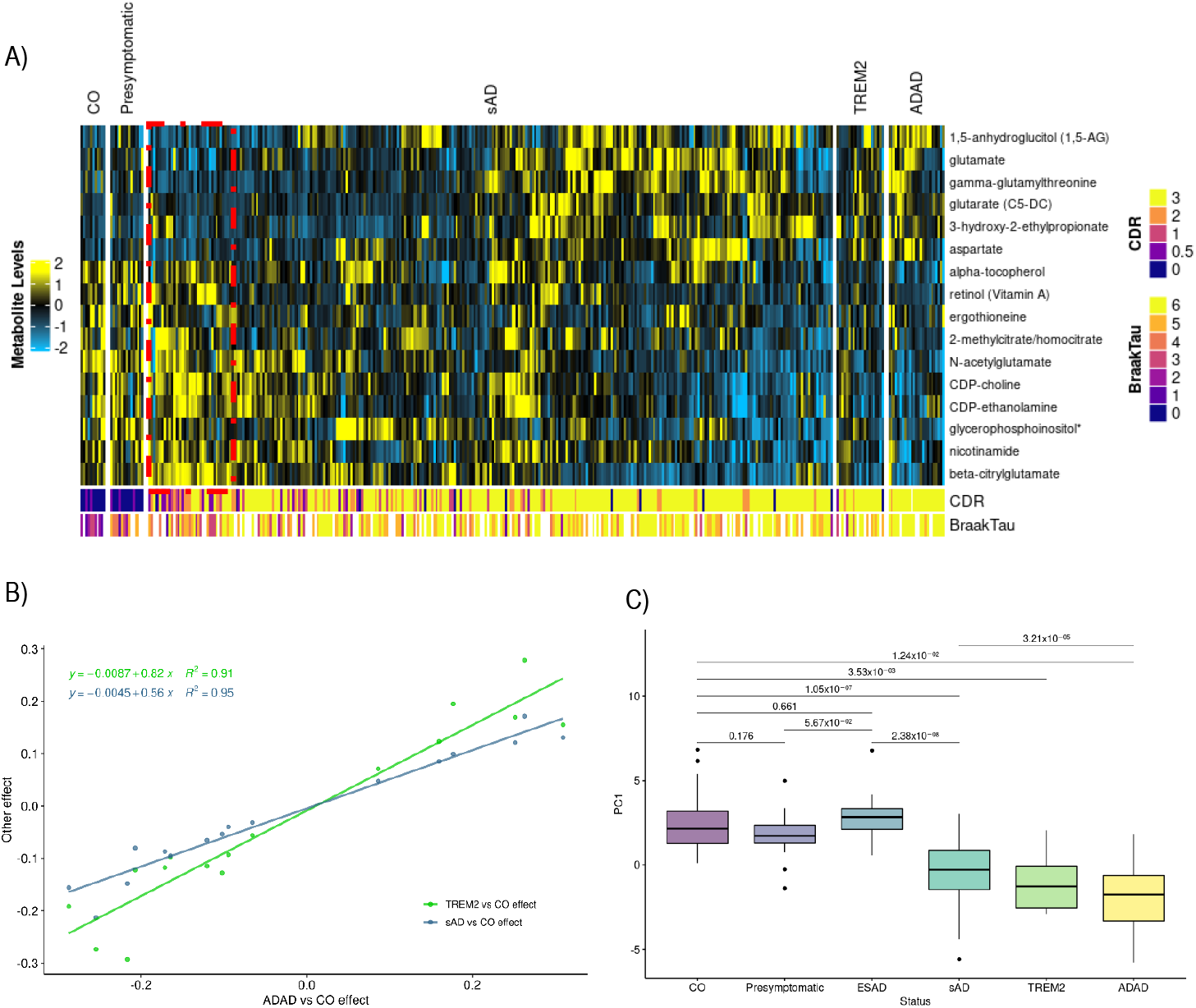
Metabolic profile consisting of 16 metabolites which passed FDR correction in ADAD vs CO and were at least nominally significant in sAD vs CO and TREM2 vs CO. A) Heatmap showing relative abundance for each metabolite in the profile. Participants are divided by disease status group: healthy controls (CO), neuropathology but no cognitive impairment (Presymptomatic), sporadic AD (sAD), carriers of TREM2 risk variants (TREM2) and carriers of Mendelian mutations (ADAD). The 30 Early-Stage AD (ESAD) individuals identified by hierarchical clustering are indicated within the red box. Annotations show Clinical Dementia Rating and Braak scores for Tau accumulation. B) Comparison of effects for the 16 metabolites in each model. The x-axis shows the effect of each metabolite in the ADAD vs CO model, while the y-axis shows the effects in the sAD vs CO (blue) and TREM2 vs CO (red) models. C) Boxplot showing distribution of the first principal component for the 16-metabolite profile among each of the status groups.

Among the 16 common metabolites across the three genetic groups, four were also associated with AAD in a linear regression corrected for sex and PMI: 1,5-anhydroglucitol (1,5-AG), glycerophosphoinositol, N-acetylglutamate, and retinol (vitamin A).

We then calculated the first principal component for these 16 metabolites to generate an “eigenmetabolite” representing the metabolic profile for each individual^42^. The eigenmetabolite was found to be associated with disease duration in sAD (*p* = 1.86×10^-02^), as well as Braak Tau stage (*p* = 4.17×10^-11^) and CDR (*p* = 4.23×10^-13^) in the entire cohort, with a lower eigenmetabolite value being associated with longer duration, higher Braak stage, and higher CDR. Eigenmetabolite values were significantly different between status groups, with ADAD, TREM2, and sAD having significantly lower eigenmetabolite values than CO and ADAD having significantly lower values than sAD **(Figure 3C**). To validate these observations, 15 metabolites with available data for the ROSMAP Metabolon dataset (all except 3-hydroxy-2-ethylpropionate) were used to generate an eigenmetabolite profile for the ROSMAP participants. The eigenmetabolite was again associated with disease duration (*p* = 2.68×10^-02^), but was not associated with Braak tau stage (*p* = 0.38). Eigenmetabolite values were not significantly associated with sAD in the neuropathological categorization (*p =* 0.91), but were associated with consensus clinical diagnosis, with lower values observed in AD participants (*p =* 2.73×10^-3^) (**Figure 4**).

**Figure 4.**
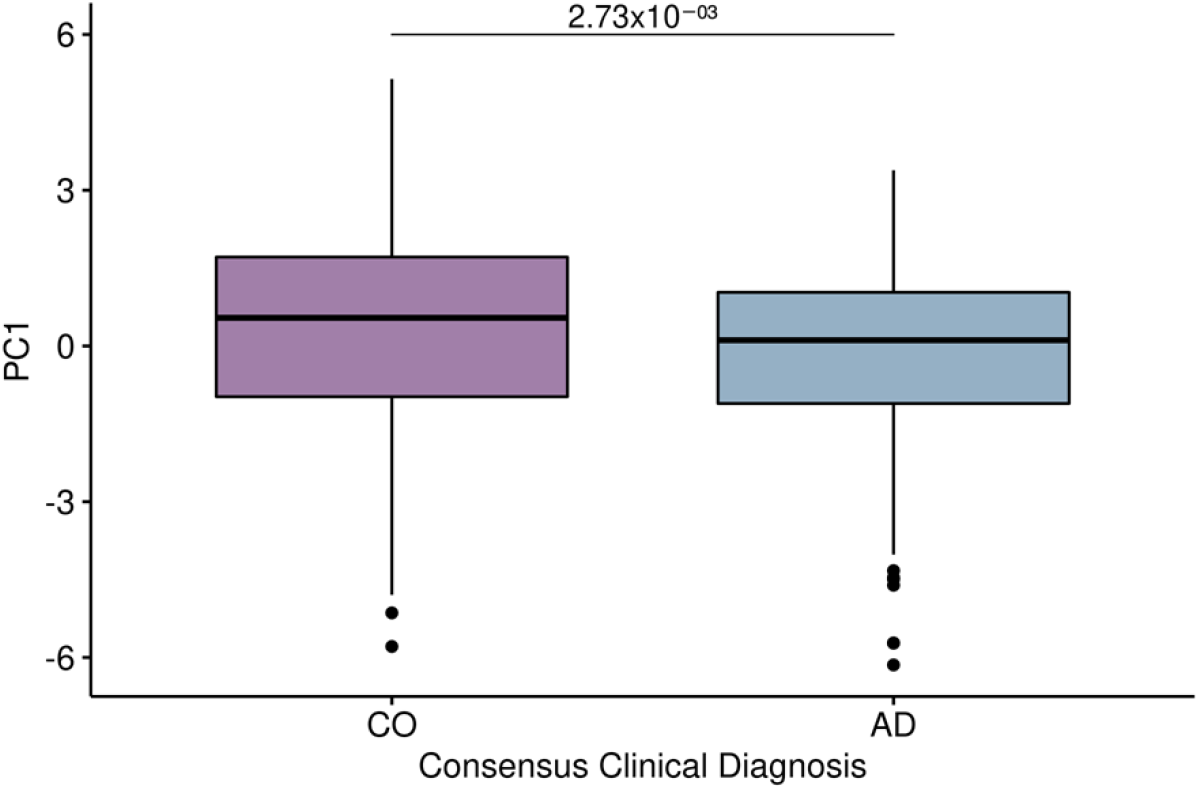
Distribution of metabolic eigenmetabolite profile between clinical diagnosis groups calculated with 15 metabolites on ROSMAP Metabolon data. 3-hydroxy-2-ethylpropionate was not included because it did not pass quality control in the ROSMAP dataset.

To visualize levels of the 16 metabolites between status groups in the WUSM cohort, a heatmap was generated with the scaled and imputed metabolite readings from Metabolon (**Figure 3A**). A group of 30 sAD individuals, identified by hierarchical clustering, with metabolite abundance profiles not significantly different from the control group was selected for further analysis. For these individuals, the eigenmetabolite was not significantly different from the control group in a logistic regression correcting for AAD, sex, and PMI (*p* = 0.66). These individuals were classified as Early-Stage AD (ESAD) after further logistic regression analysis of CDR, Braak tau, and disease duration, correcting for the same variables. The ESAD group showed lower CDR (ESAD = 1.67±1.09, sAD = 2.56±0.84, effect = -0.83, *p* = 8.02×10^-6^) and Braak Tau (ESAD = 4.05±1.36, sAD = 5.30±1.06, effect = -0.66, *p* = 1.22×10^-4^) compared to sAD. The ESAD individuals also showed a shorter disease duration (ESAD = 7.97±5.33, sAD = 10.03±4.61, effect = -0.09, *p* = 4.42×10^-2^) than the rest of the sAD individuals.

To explore the relationship between this metabolic profile and disease progression, presymptomatic individuals were also considered. The eigenmetabolite values for the presymptomatic group were not significantly different from the CO and ESAD groups (*p* = 0.18 and *p* = 0.06, respectively) **(Figure 3C)**. As expected, the presymptomatic status group showed significantly lower CDR (effect = -7.52, *p* = 2.47×10^-03^) than the ESAD group. However, the presymptomatic group did not differ from ESAD in the Braak Tau stage (*p* = 0.92).

The super pathways associated with these 16 metabolites were mostly related to amino acid metabolism (glutamate, arginine, lysine, glutathione, histidine, tryptophan), but phospholipid and vitamin pathways were also identified. CDP-ethanolamine, CDP-choline, and glycerophosphoinositol were associated with phospholipid metabolism, while α-tocopherol (vitamin E), retinol (vitamin A), and nicotinamide (vitamin B3) were components of vitamin metabolism.

Considering that three of the metabolites in the eigenmetabolite profile were vitamins: retinol (vitamin A), α-tocopherol (vitamin E), and nicotinamide (vitamin B3), we also investigated participants who had taken vitamin supplements to rule out any confounding association. Within the five years preceding their deaths, 132 individuals took vitamin E supplements, 87 individuals took vitamin A, and 101 individuals took vitamin B3 **(Supplementary Table 9)**. When regressions were repeated excluding individuals who took vitamin E supplements, we observed that all of the associations remained significant in TREM2 (*p =* 3.9×10^-04^), ADAD (*p =* 2.9×10^-05^), and sAD (*p =* 2.9×10^-04^). Similarly, excluding participants taking vitamin A supplements did not affect the association of retinol with any genetic group (ADAD *p =* 6.11×10^-04^, AD *p =* 1.40×10^-02^, TREM2 *p =* 4.75×10^-02^). When participants who took vitamin B3 were excluded, ADAD and TREM2 were still nominally associated with nicotinamide (ADAD *p =* 9.4×10^-03^, TREM2 *p =* 3.7×10^-02^), however, sAD was no longer associated (*p =* 0.11). Removing nicotinamide from the eigenmetabolite did not affect the eigenmetabolite association with disease duration (effect = -0.052, *p* = 1.75×10^-02^), CDR (effect = -0.65, *p* = 4.49×10^-14^), or Braak tau (effect = -0.57, *p* = 6.75×10^-12^).

### Web Browser

The browser facilitates exploration of our analyses and further investigation into individual metabolites by integrating metadata with visualizations of our results. The browser has two main pages, or tabs. The first displays a table including metadata on each metabolite that passed our QC process, along with its effect, *p*-value, and *q*-value for each comparison discussed here. The table allows the user to select a metabolite, which displays the distribution of the selected metabolite’s readings across disease statuses. Links are also provided to the PubChem (https://pubchem.ncbi.nlm.nih.gov/)^67^ and Human Metabolome Database (www.hmdb.ca)^46,68^ webpages if the IDs are available. The second tab displays volcano plots for each regression, with *q*-values less than 0.05 highlighted. Again, the user may select a metabolite on the volcano plot to view its reading distributions among statuses and display further information and links.

## DISCUSSION

In this study, we have performed a metabolomics analysis of parietal cortical tissue from participants of the DIAN and Knight ADRC cohorts. We have characterized the metabolomic profile of three genetically defined AD subgroups including ADAD, carriers of risk variants in TREM2, and sAD. We have analyzed the detailed phenotypic information available for these brains, including genetic risk factors and clinical, pharmacological, and neuropathological variables.

We found a significantly different metabolic profile in ADAD patients from that of healthy individuals, with 131 significant metabolites linked to ADAD, altering multiple pathways including the γ-glutamyl cycle, tRNA charging, and aminoacyl-tRNA biosynthesis (**Supplementary Tables 7 and 8**). The parietal cortex of ADAD individuals has been reported to have a higher burden of neurofibrillary tangles (NFT) than that of sAD individuals^69^. Accordingly, the metabolic profiles of TREM2 and sAD showed fewer differences than ADAD from that of healthy individuals, and of the two only sAD showed metabolite differences unique to its category (tryptophan betaine and N-acetylputrescine). Tryptophan betaine is an N-methylated form of tryptophan, which is the serotonin precursor and has been found de-regulated in MCI-AD^70^. N-acetylputrescine is an acetyl-CoA-ated putrescine and a GABA precursor that was found to build up in stable MCI but not in AD, where putrescine is preferentially metabolized to other polyamines^71^. We also observe depleted N-acetylputrescine levels in sAD in our data (**Supplementary Figure 3**) which supports previously reported findings of lower GABA levels in AD^72^.

Among the metabolites identified as differentially abundant in at least one group were BCG, α-tocopherol, and ergothioneine. Each of these metabolites showed lower concentrations in an AD subgroup compared to control. BCG acts as an iron carrier to activate aconitase activity^73^. We observed that BCG had the lowest abundance in ADAD, again followed by TREM2 and sAD. This observation could be associated with a lower activation of aconitase and lower energetic metabolism. BCG is also a component of glutamate metabolism, and BCG levels can be increased by the selective serotonin reuptake inhibitor (SSRI) fluoxetine^65^. BCG levels in our cohort were not significantly associated with fluoxetine administration, indicating that the association of BCG and AD in the three genetic groups is not driven by fluoxetine usage. Within the vitamin pathway, α-tocopherol (vitamin E) was differentially abundant in both TREM2 and ADAD vs CO, and vitamin E supplementation in participants did not affect this association. Vitamin E is a powerful antioxidant that aids the immune system and keeps blood clots from forming^74–76^. This finding complements our observation of lower BCG in sAD cases. Reduction of aconitase activity due to oxidative stress in aging could be exacerbated in AD by lower levels of antioxidants such as vitamin E. This could lead to less energetic metabolism activation overall. Ergothioneine was also observed at lower levels in sAD and ADAD cases compared to controls. Ergothioneine is a naturally occurring amino acid and thiourea derivative of histidine produced by fungi, which has antioxidant and anti-inflammatory properties^77,78^. The main source of ergothioneine in humans is diet; it accumulates in erythrocytes and crosses the blood–brain barrier^79^. However, its physiological role in humans is not known. Ergothioneine blood levels in humans decline with age and decline faster in individuals with cognitive impairment compared to age-matched individuals with no cognitive impairment^80^. In mice treated with intracerebroventricular injection of Aβ1-40, ergothioneine protected against loss of memory and learning abilities^81^.

In addition to BCG, α-tocopherol, and ergothioneine, we identified eight metabolites in the Vitamins pathway that were significant after FDR correction in ADAD vs healthy individuals and significant before correction in the AD vs CO, TREM2 vs CO, and ROSMAP brain analyses. Four of these (2-aminoadipate, serotonin, tryptophan and tyrosine) were also significant in the serum meta-analysis and are important neurotransmitters.

Neurotransmitters, especially serotonin, have been shown to play a role in processing APP and reducing generation of Aβ_42_ through activation of the ERK signaling cascade^82^. In our study, serotonin levels were significantly decreased in sAD and ADAD compared to control, and nominally decreased in TREM2. This effect was replicated in independent datasets of both serum and DLPFC tissue. SSRIs, which increase serotonin levels in the brain, show promise for reduction of Aβ accumulation in both the brain and CSF. Studies in APP/PS1 transgenic mice showed that the SSRI citalopram caused a 50% reduction in brain amyloid plaque load, and escitalopram, citalopram’s S-isomer, reduced interstitial fluid Aβ by 25%^82,83^. A controlled clinical trial of cognitively normal adults showed that escitalopram could decrease CSF Aβ_42_ levels in humans, with a difference of 11.1% between the control and treatment groups^84^. Our results corroborate the association of low serotonin with AD, and suggest that this effect, and potentially the benefit of serotonin modulation via SSRIs, spans all three genetic groups.

Our discovery dataset identified a set of 16 metabolites whose first principal component, or eigenmetabolite, was distinct between the AD groups and healthy individuals, and between sAD and ADAD. The effects for these metabolites were greatest in ADAD, followed by TREM2 and sAD. The eigenmetabolite was additionally associated with CDR, Braak tau stage, and disease duration. The association with disease duration was validated in an independent dataset using 15 of the 16 metabolites. We also evaluated the performance of these 16 metabolites in a group of presymptomatic individuals and observed that this group showed a similar profile to that of healthy individuals. In addition, we identified a set of sAD cases (Early-Stage AD/ESAD) with a metabolic profile close to that of the healthy individuals. Further examination of these individuals revealed significantly lower CDR and Braak Tau scores than the rest of sAD individuals. The presymptomatic and ESAD groups showed no significant difference in the metabolic eigenmetabolite or Braak tau but significantly different CDR. Furthermore, ADAD individuals, known to have an earlier age at onset and higher NFT burden, showed a greater effect for these metabolites. Together, these observations suggest that the metabolic profile could be driven by tau pathology and implicated in disease duration.

Spermidine was negatively associated with age in sAD and showed increased levels in sAD in the ADNIGO/2 dataset compared to control (**Supplementary Tables 5 and 6**). However, we did not find it associated with any genetic group in the Knight ADRC dataset. Putrescine was significantly decreased in sAD before correction (**Supplementary Table 5**). N-acetylputrescine, a putrescine derivative, was significantly decreased in the sAD group compared to control and significantly decreased before FDR correction in TREM2. Schroeder, et al. (2021) found that polyamines, particularly spermidine, improved cognition in mice by enhancing mitochondrial function through hypusination of eukaryotic translation initiation factor 5A (eIF5A)^85^. Liang, et al. (2021) also showed that eIF5a activity decreased with age in fly models, and that spermidine supplementation could improve mitochondrial function^86^.

This study identified differences in metabolite abundance both specific to and common between genetically defined AD subgroups. We replicated our main findings in three independent datasets. Differences in the levels of common metabolites allow us to generate a metabolic profile associated with disease duration, CDR, and Braak tau stage and that further identified a subset of AD cases with a profile similar to CO (ESAD). Metabolomics of the brain can identify metabolic signatures specific to AD genetic subgroups. These metabolites may support the creation of “metabolomics scores” to assess disease status. Limitations of our study include the sample size of some of the genetic groups, e.g. TREM2. As such, in future studies we would like to increase the sample size of our brain-sourced dataset. We were unable to find associations with Braak Aβ stage possibly because scores are unavailable for many participants. Direct replication of our results in ADAD individuals was unachievable due to a lack of independent ADAD datasets. Unlike previous studies, we did not find significant associations between *APOE ε4* carriers and non-carriers in cases of sporadic AD. In addition, our ability to replicate findings in other tissues, such as blood serum, was possibly limited due to different platforms used by other studies. In future studies we will extend our analysis to serum metabolomics data and seek replication of our findings to facilitate further identification of novel biomarkers for AD.

## Data Availability

Metabolomics data from the Knight ADRC donors generated for this study are available at the NIAGADS and can be accessed at https://www.niagads.org/knight-adrc-collection. Data generated from the DIAN cohort can be requested at https://dian.wustl.edu/our-research/for-investigators/diantu-investigator-resources/dian-tu-biospecimen-request-form/. We have accessed data from the ADNI (https://adni.loni.usc.edu, accessed 18 December, 2020), and ROSMAP (https://synapse.org/#!Synapse:syn26007829, accessed 18 December, 2020 and https://synapse.org/#!Synapse:syn26007830, accessed 30 July, 2021). Additional phenotypic data for the ROSMAP studies is available through the Rush AD Center Resource Sharing Hub (https://www.radc.rush.edu).

https://adni.loni.usc.edu

https://synapse.org/#!Synapse:syn26007829

https://synapse.org/#!Synapse:syn26007830

https://www.radc.rush.edu

## Funding

This work was possible thanks to the following governmental grants from the National institute of Health: NIA R01AG057777, RO1AG057777-02S1, K99AG061281, P30AG066444, P01AGO26276, NINDS R01NS118146 (BAB), R01AG044546 (CC), P01AG003991 (CC, JCM), RF1AG053303 (CC), RF1AG058501 (CC), U01AG058922 (CC), and the Chan Zuckerberg Initiative (CZI). O.H. is an Archer Foundation Research Scientist.

This work was supported by access to equipment made possible by the Hope Center for Neurological Disorders, the NeuroGenomics and Informatics Center (NGI: https://neurogenomics.wustl.edu/) and the Departments of Neurology and Psychiatry at Washington University School of Medicine.

## Acknowledgements

We thank contributors who collected samples used in this study and patients and their families, whose help and participation made this work possible.

## Dominantly Inherited Alzheimer Network (DIAN) resources

Data collection and sharing for this project was supported by The Dominantly Inherited Alzheimer Network (DIAN, U19AG032438) funded by the National Institute on Aging (NIA),the Alzheimer’s Association (SG-20-690363-DIAN), the German Center for Neurodegenerative Diseases (DZNE), Raul Carrea Institute for Neurological Research (FLENI), Partial support by the Research and Development Grants for Dementia from Japan Agency for Medical Research and Development, AMED, and the Korea Health Technology R&D Project through the Korea Health Industry Development Institute (KHIDI), Spanish Institute of Health Carlos III (ISCIII), Canadian Institutes of Health Research (CIHR), Canadian Consortium of Neurodegeneration and Aging, Brain Canada Foundation, and Fonds de Recherche du Québec – Santé. This manuscript has been reviewed by DIAN Study investigators for scientific content and consistency of data interpretation with previous DIAN Study publications. We acknowledge the altruism of the participants and their families and contributions of the DIAN research and support staff at each of the participating sites for their contributions to this study.

## DIAN Study Group

Sarah Adams, Ricardo Allegri, Aki Araki, Nicolas Barthelemy, Randall Bateman, Jacob Bechara,Tammie Benzinger, Sarah Berman, Courtney Bodge, Susan Brandon, William (Bill) Brooks, Jared Brosch, Jill Buck, Virginia Buckles, Kathleen Carter, Lisa Cash, Charlie Chen, Jasmeer Chhatwal, Patricio Chrem, Jasmin Chua, Helena Chui, Carlos Cruchaga, Gregory S Day, Chrismary De La Cruz, Darcy Denner, Anna Diffenbacher, Aylin Dincer, Tamara Donahue, Jane Douglas, Duc Duong, Noelia Egido, Bianca Esposito, Anne Fagan, Marty Farlow, Becca Feldman, Colleen Fitzpatrick, Shaney Flores, Nick Fox, Erin Franklin, Nelly Friedrichsen, Hisako Fujii, Samantha Gardener, Bernardino Ghetti, Alison Goate, Sarah Goldberg, Jill Goldman, Alyssa Gonzalez, Brian Gordon, Susanne Gräber-Sultan, Neill Graff-Radford, Morgan Graham, Julia Gray, Emily Gremminger, Miguel Grilo, Alex Groves, Christian Haass, Lisa Häsler, Jason Hassenstab, Cortaiga Hellm, Elizabeth Herries, Laura Hoechst-Swisher, Anna Hofmann, David oltzman, Russ Hornbeck, Yakushev Igor, Ryoko Ihara, Takeshi Ikeuchi, Snezana Ikonomovic, Kenji Ishii, Clifford Jack, Gina Jerome, Erik Johnson, Mathias Jucker, Celeste Karch, Stephan Käser, Kensaku Kasuga, Sarah Keefe, William (Bill) Klunk, Robert Koeppe, Deb Koudelis, Elke Kuder-Buletta, Christoph Laske, Allan Levey, Johannes Levin, Yan Li, Oscar Lopez, Jacob Marsh, Rita Martinez, Ralph Martins, Neal Scott Mason, Colin Masters, Kwasi Mawuenyega, Austin McCullough, Eric McDade, Arlene Mejia, Estrella Morenas-Rodriguez, John Morris, James MountzMD, Cath Mummery, Neelesh Nadkarni, Akemi Nagamatsu, Katie Neimeyer, Yoshiki Niimi, James Noble, Joanne Norton, Brigitte Nuscher, Antoinette O’Connor, Ulricke Obermüller, Riddhi Patira, Richard Perrin, Lingyan Ping, Oliver Preische, Alan Renton, John Ringman, Stephen Salloway, Peter Schofield, Michio Senda, Nick Seyfried, Kristine Shady, Hiroyuki Shimada, Wendy Sigurdson, Jennifer Smith, Lori Smith, Beth Snitz, Hamid Sohrabi, Sochenda Stephens, Kevin Taddei, Sarah Thompson, Jonathan Vöglein, Peter Wang, Qing Wang, Elise Weamer, Chengjie Xiong, Jinbin Xu, Xiong Xu

## Alzheimer’s Disease Metabolomics Consortium (ADMC)

The results published here are in whole or in part based on data obtained from the AD Knowledge Portal (https://adknowledgeportal.org). Metabolomics data is provided by the Alzheimer’s Disease Metabolomics Consortium (ADMC) and funded wholly or in part by the following grants and supplements thereto: NIA R01AG046171, RF1AG051550, 3U01AG024904-09S4, RF1AG057452, R01AG059093, RF1AG058942, U01AG061359, U19AG063744 and FNIH: #DAOU16AMPA awarded to Dr. Kaddurah-Daouk at Duke University in partnership with a large number of academic institutions. As such, the investigators within the ADMC, not listed specifically in this publication’s author’s list, provided data along with its pre-processing and prepared it for analysis, but did not participate in analysis or writing of this manuscript. A complete listing of ADMC investigators can be found at: https://sites.duke.edu/adnimetab/team/.

## ADNI

Data collection and sharing for this project was funded by the Alzheimer’s Disease Neuroimaging Initiative (ADNI) (National Institutes of Health Grant U01 AG024904) and DOD ADNI (Department of Defense award number W81XWH-12-2-0012). ADNI is funded by the National Institute on Aging, the National Institute of Biomedical Imaging and Bioengineering, and through generous contributions from the following: AbbVie, Alzheimer’s Association; Alzheimer’s Drug Discovery Foundation; Araclon Biotech; BioClinica, Inc.; Biogen; Bristol-Myers Squibb Company; CereSpir, Inc.; Cogstate; Eisai Inc.; Elan Pharmaceuticals, Inc.; Eli Lilly and Company; EuroImmun; F. Hoffmann-La Roche Ltd and its affiliated company Genentech, Inc.; Fujirebio; GE Healthcare; IXICO Ltd.;Janssen Alzheimer Immunotherapy Research & Development, LLC.; Johnson & Johnson Pharmaceutical Research & Development LLC.; Lumosity; Lundbeck; Merck & Co., Inc.;Meso Scale Diagnostics, LLC.; NeuroRx Research; Neurotrack Technologies; Novartis Pharmaceuticals Corporation; Pfizer Inc.; Piramal Imaging; Servier; Takeda Pharmaceutical Company; and Transition Therapeutics. The Canadian Institutes of Health Research is providing funds to support ADNI clinical sites in Canada. Private sector contributions are facilitated by the Foundation for the National Institutes of Health (www.fnih.org). The grantee organization is the Northern California Institute for Research and Education, and the study is coordinated by the Alzheimer’s Therapeutic Research Institute at the University of Southern California. ADNI data are disseminated by the Laboratory for Neuro Imaging at the University of Southern California.

## ROSMAP

Study data were provided by the Rush Alzheimer’s Disease Center, Rush University Medical Center, Chicago. Data collection was supported through funding by NIA grants P30AG10161 (ROS), R01AG15819 (ROSMAP; genomics and RNAseq), R01AG17917 (MAP), R01AG30146, R01AG36042 (5hC methylation, ATACseq), RC2AG036547 (H3K9Ac), R01AG36836 (RNAseq), R01AG48015 (monocyte RNAseq) RF1AG57473 (single nucleus RNAseq), U01AG32984 (genomic and whole exome sequencing), U01AG46152 (ROSMAP AMP-AD, targeted proteomics), U01AG46161(TMT proteomics), U01AG61356 (whole genome sequencing, targeted proteomics, ROSMAP AMP-AD), the Illinois Department of Public Health (ROSMAP), and the Translational Genomics Research Institute (genomic). Additional phenotypic data can be requested at www.radc.rush.edu. Study data were provided through NIA grant 3R01AG046171-02S2 awarded to Rima Kaddurah-Daouk at Duke University, based on specimens provided by the Rush Alzheimer’s Disease Center, Rush University Medical Center, Chicago, where data collection was supported through funding by NIA grants P30AG10161, R01AG15819, R01AG17917, R01AG30146, R01AG36836, U01AG32984, U01AG46152, the Illinois Department of Public Health, and the Translational Genomics Research Institute.

## Additional Acknowledgements

We would like to pay our gratitude and respects to our friend and collaborator, Jorge Bahena. Jorge was a remarkable scientist and respected colleague who earned his master’s degree in biostatistics from Washington University School of Medicine, and passed away in October 2021 as a doctoral student at Vanderbilt University. His valuable contributions to this and many other endeavors will not be forgotten.

## Financial disclosures

CC receives research support from: Biogen, EISAI, Alector and Parabon. The funders of the study had no role in the collection, analysis, or interpretation of data; in the writing of the report; or in the decision to submit the paper for publication. CC is a member of the advisory board of Vivid genetics, Halia Therapeutics and ADx Healthcare.

## Notes

### Author Declarations

IRB of Washington University School of Medicine in St. Louis gave ethical approval for this work.

