## Supplementary Figures for "Metabolomic and lipidomic signatures in autosomal dominant and late-onset Alzheimer disease brains" (from another version of this preprint: version 1, 2021-11-05)

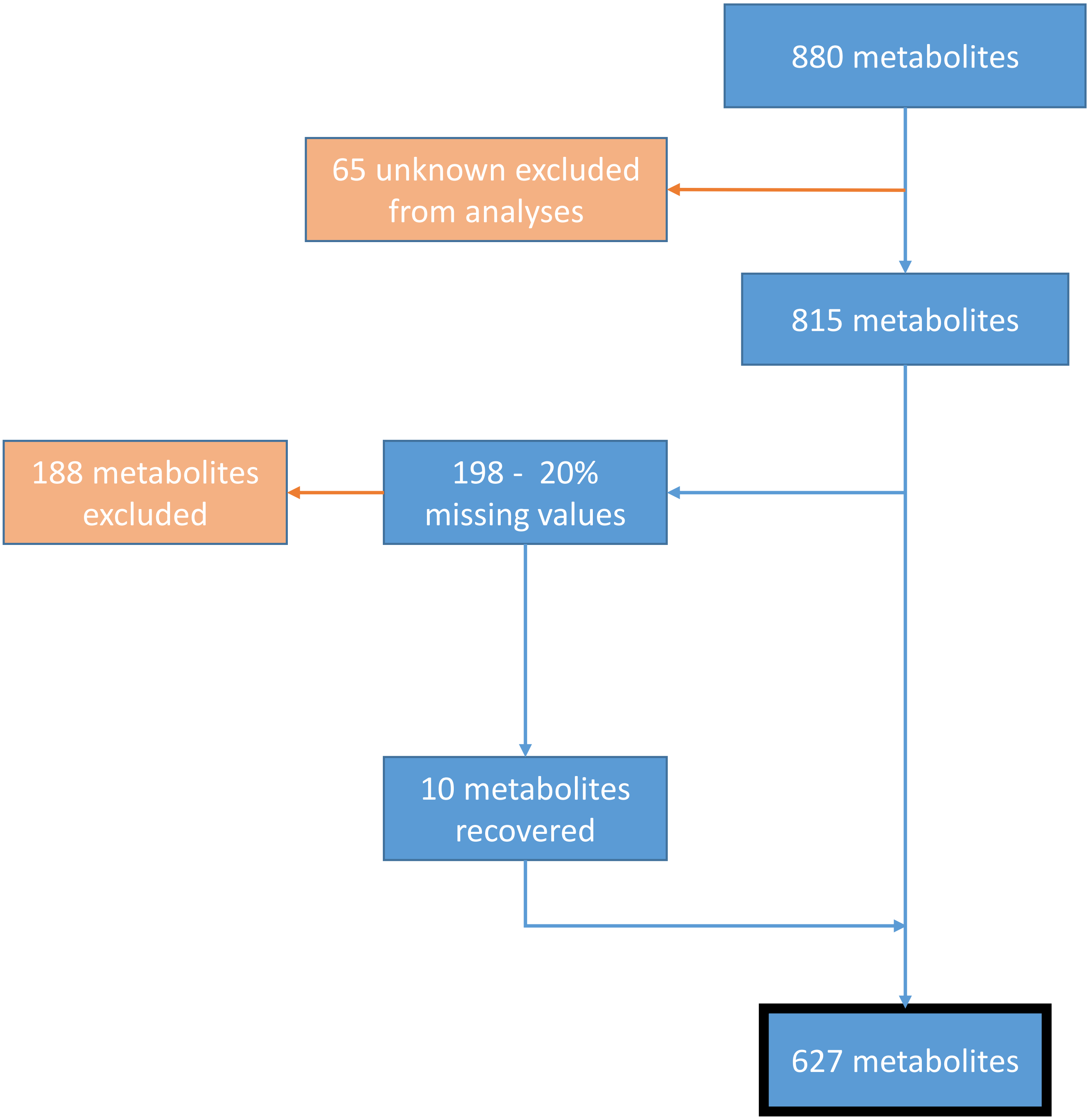


Supplementary Figure 1. Metabolite QC pipeline. Metabolites with unknown structural identity were excluded from further analysis, as well as metabolites with values missing in 20% or more of samples. Metabolites with high missingness were recovered if their missingness was significant between disease status groups, and corresponded to a significant difference in metabolite levels between those groups.

A)

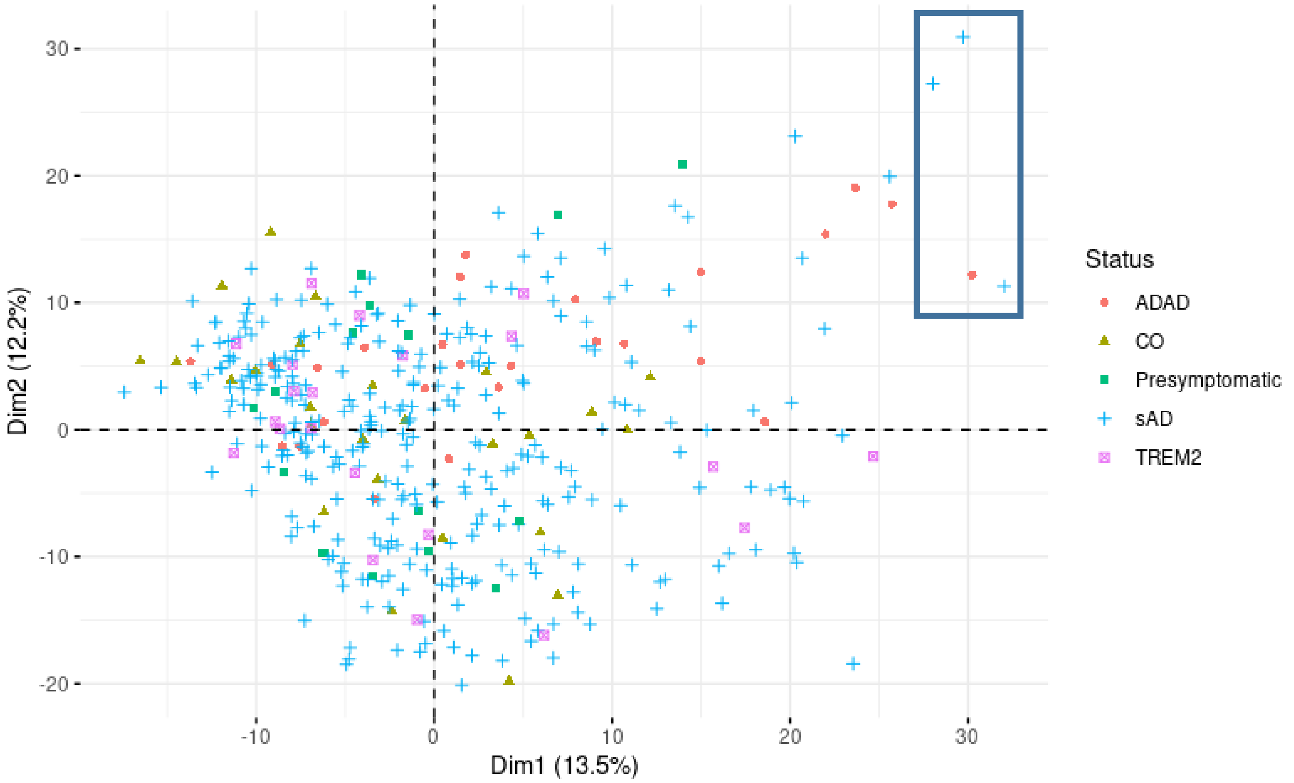


B)

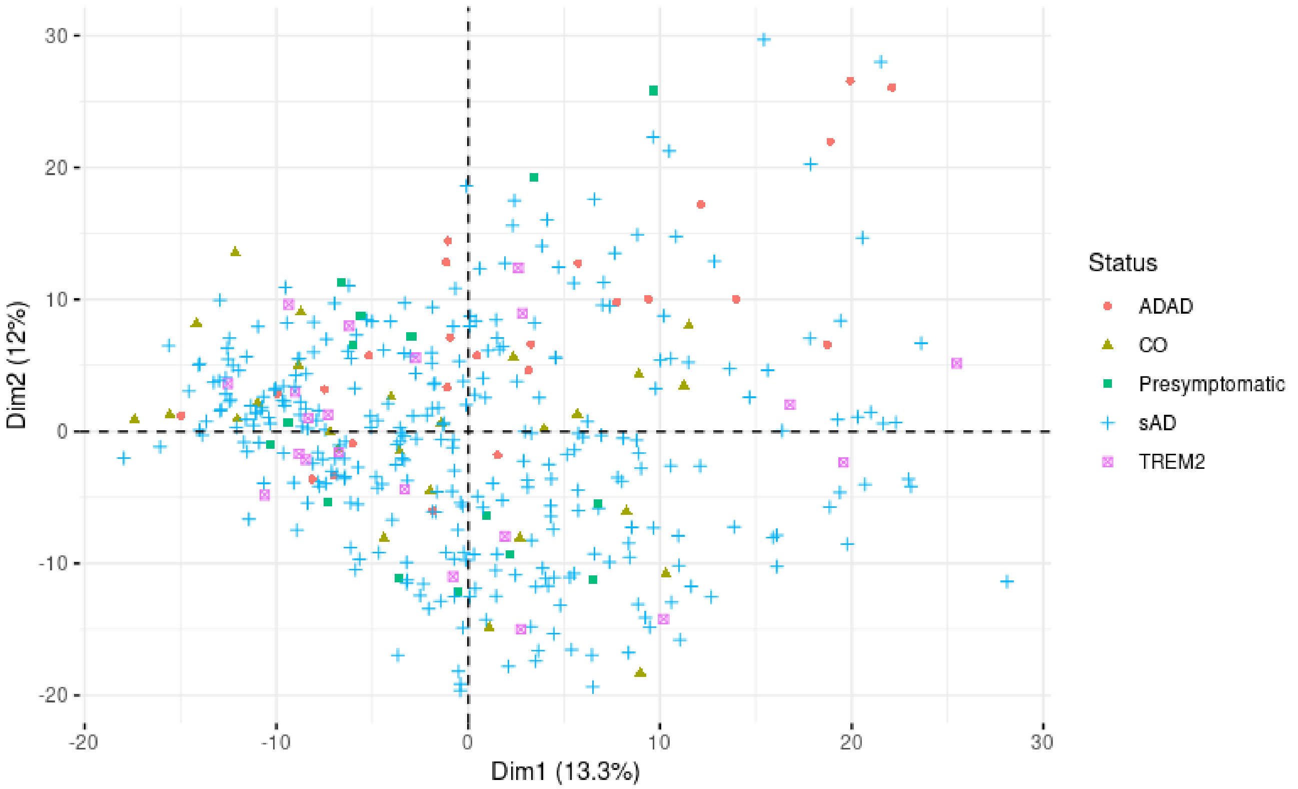


Supplementary Figure 2. PCA plot for individuals A) Before sample quality control; PCs were calculated with the 627 metabolites that passed QC. Outlier samples which were dropped are indicated inside the rectangle. B) After sample QC.

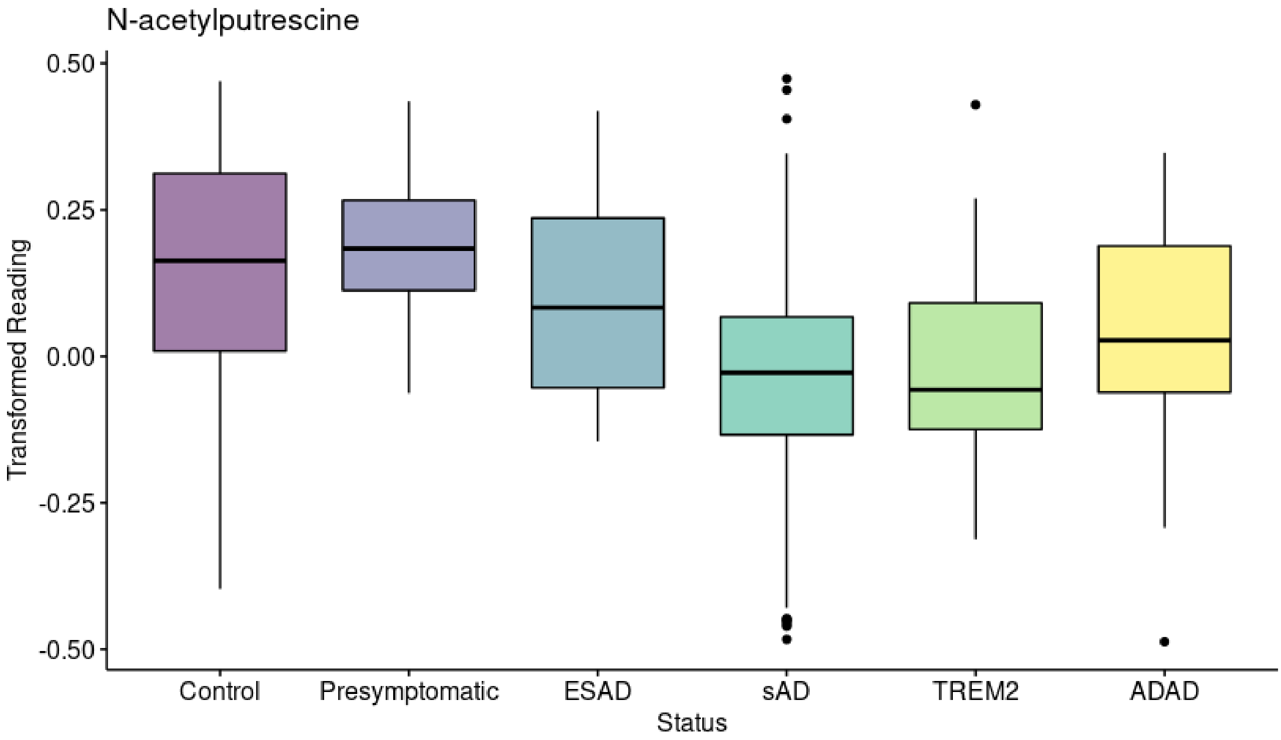


Supplementary Figure 3. Boxplot showing distribution of N-acetylputrescine levels across status groups. Levels remain higher in Presymptomatic and ESAD individuals, dropping significantly in the sAD group.
